# The clustering of multiple health and lifestyle behaviours among Swedish adolescents: A person-oriented analysis

**DOI:** 10.1101/2023.02.09.23285615

**Authors:** Kenisha Russell Jonsson, Maria Corell, Petra Löfstedt, Nicholas Kofi Adjei

## Abstract

**Background:** Knowledge of the distribution, prevalence, and clustering of multiple health and lifestyle related behaviours (HLBs) among adolescents can inform the development of more effective health-promoting policies and interventions. We therefore assessed the clustering of multiple HLBs among 11, 13 and 15-year-old Swedish adolescents and examined the socioeconomic and demographic correlates of each cluster.

**Methods:** We used data from the 2017/2018 Swedish Health Behaviour in School-aged Children (HBSC) study to conduct sex and age-stratified latent class analysis (LCA). The LCA was based on five HLBs: eating behaviour and habits (EBH), physical activity (PA), tobacco usage (TU), alcohol consumption (AC) and sleeping habits and patterns (SHPs). Multinomial logistic regression models were used to assess the associations between the identified clusters and the socioeconomic and demographic characteristics of adolescents and their parents.

**Results:** Health behaviours varied by age and sex. Four distinct clusters were identified based on sex: cluster 1 (Mixed eating behaviours and habits, physical activity and low alcohol consumption), cluster 2 Healthy lifestyle behaviours), cluster 3 (Unhealthy lifestyle behaviours), and cluster 4 (Breakfast, low alcohol consumption and tobacco usage). In the age-stratified analyses, three clusters were identified: cluster 1 (Unhealthy lifestyle behaviours), cluster 2 (Moderately healthy lifestyle behaviours) and cluster 3 (Healthy lifestyle behaviours). The multinomial analysis showed that sex, age, family situation and perceived family wealth were strong predictors of health behaviours. In particular, unhealthy behaviours showed the most frequent associations with socioeconomic disadvantage, having a migrant background and living in reconstructed families or single parent households.

**Conclusions:** Health behaviours vary significantly based on socioeconomic and demographic factors. Targeted policy and interventions programmes can effectively improve HLBs among vulnerable and at-risk adolescents.

## INTRODUCTION

Health and lifestyle behaviours (HLBs) in adolescence, such as eating behaviour and habits (EBH), physical activity (PA), tobacco usage (TU), alcohol consumption (AC) and sleeping habits and patterns (SHP) may have both short and long-term health consequences [1-5]. In Sweden, there are public health concerns about the impact of HLBs on adolescents physical and mental health [6, 7]. Adolescence is a critical developmental stage and an important time where HLBs and inequalities are shaped [8-12]. Prior evidence suggests that HLBs formed during adolescence persist into adulthood [8-12], and engagement in unhealthy lifestyle behaviours may increase the immediate risks of poor physical health and mental health problems [1, 4, 6, 13-15], as well as increase the risk of morbidity and mortality in adulthood [2, 3, 16].

Poor HLBs have been linked to significant increases in non-communicable diseases (NCDs) such as overweight and obesity, diabetes, cancers and heart diseases [17]. NCDs are known contributors to the burden of disease and premature death among adults in Sweden [13], resulting in significant costs to the health and care systems [18]. Moreover, research has shown that EBH, PA, TU, AC and SHP tend to co-occur in complex ways [19, 20], and that the combination of these behaviours may have more conducive or detrimental effects on an individual’s health, than the effects of single HLBs [21]. This suggests that the effects of HLBs may be multiplicative and/or cumulative rather than additive [21]. Previous studies from high income countries demonstrated that HLBs tend to cluster among people with similar socioeconomic and demographic circumstances [20-24], with those in lower socioeconomic circumstances being associated with less healthy behaviours [20, 22-25].

Socioeconomic inequalities in health have been identified in Sweden [26, 27], suggesting that patterns of health behaviours may cluster differently based on socioeconomic and demographic factors. Therefore, understanding how multiple HLBs cluster is crucial in order to develop appropriate public health strategies and interventions [19, 28]. However, the clustering of health behaviours among adolescents, and the social and economic determinants of multiple HLBs among adolescents is understudied [24, 42], leading to a knowledge gap. This study will therefore contribute to the literature by assessing the patterning of multiple HLBs in the Swedish context.

### Research aims

The study aimed to assess the clustering of five health and lifestyle behaviours ((eating behaviour and habits (EBH), physical activity (PA), tobacco usage (TU), alcohol consumption (AC) and sleeping habits and patterns (SHP)) among Swedish adolescents aged 11, 13 and 15-stratified by age and sex. The second aim was to investigate the associations between the identified clusters and socioeconomic and demographic characteristics of adolescents and their parents (i.e., sex, age, migrant background, family situation, perceived family wealth and family affluence).

## METHODS

### Data and sample

The analyses were based on a nationally representative, cross-sectional sample of adolescents from the 2017/2018 Swedish component of the Health Behaviour in School-Aged Children (HBSC) survey. It is a World Health Organization (WHO) collaborative study [29], conducted by the Public Health Agency of Sweden and Statistics Sweden. The Swedish HBSC participants were selected based on a two-stage cluster design. First, a nationally representative sample of 450 schools were selected, and in the second stage, a single class from each school (5^th^ graders ≈ 11 year olds, 7^th^ graders ≈ 13 year olds, 9 graders ≈ 1 year olds) was randomly selected to participate. All students in the selected classes were then invited to participate in the survey. The final sample included 4 215 students from 213 schools, with a response rate of 89 % in the participating schools, consisting of 2 114 girls (50.2%) and 2101 boys (49.8%).

### Health and lifestyle behaviour measures

#### Eating behaviours and habits (EBH)

We assessed EBH using seven individual items capturing the frequency and quality of food consumption among adolescents. The first two items assessed the regularity of breakfast consumption on school days and weekends/non-school days. Respondents were asked to estimate the number of weekdays/weekends they had breakfast (more than having more than a glass of milk or fruit juice). The responses provided were “every day”, “5-6 days a week”, “3-4 days a week”, “1-2 days a week”, “less than once a week” and “never”; these were dichotomised as every day vs less than every day. Four items designed to assess the consumption of dietary fibre, calcium, sweetened beverages and sugary foods were based on responses to the number of times in a week adolescents consumed each of the following: (1) fruits, (2) vegetables, (3) sugary soft drinks and; (4) sweets (including chocolate). Possible responses were: “never”, “less than once a week”, “once a week”, “two to four times a week”, “five to six times a week”, “once a day”, “more than once a day”. The consumption of fruits and vegetables were then dichotomized into “at least once a day” vs “less than once a day”. The consumption of sugary soft drinks and sugary foods were categorised as “once a week or less” vs “more than once a week”.

EBH was operationalised to ensure comparability with international HBSC reporting standards [30, 31]. The exception is sugary soft drinks and sweets, which were coded to reflect Swedish traditions of saturday candy ‘lördagsgodis’ and cosy Fridays ‘fredagsmys’, where families indulge in these dietary behaviours [32].

#### Physical activity (PA)

PA was captured with responses to the question, “how many days in the past week were you physically active for 60 minutes or more?”, and this was exemplified as any activity that increased the heart rate and breathlessness. The responses provided ranged from 0-7 days. From this, two separate measures were coded. First, *Low PA* was operationalized as respondents with PA for at least 1 hour but on less than three days during the week. The second measure, respondents engaging in PA for at least one hour per day, 7 days per week were categorised as having high PA. In accordance with the WHO physical activity recommendations [33], these items reflect low respective sufficient physical activity levels for an adolescent. In addition to information on general PA, the HBSC study also collects data on adolescents’ participation in any sports-related activities during outside of school hours i.e. during leisure time. This was assessed with the question, “Outside of school hours, how many days per week do you usually exercise in your free time so much that you get out of breath or sweat?” The possible responses ranged from 1 (every day) to 7 (never). This was dichotomised to create a measure of *Vigorous Physical Activity (VPA)* indicating adolescents engaged sports-related PA at least 4 times per week vs those engaging in sports-related PA less than 4 times per week. This was coded in line with international standards and previous usage [33, 34].

#### Tobacco Use

Tobacco usage (TU) was assessed with two questions asking “how many days (if any) have you smoked cigarettes in your life?” and “how many days (if any) have you used snus/snuff in your life?’. Possible responses ranged from 1 (never) to 7 (30 days or more)[30]. The responses from both were re-categorised into a single dichotomous measure with responses coded 1 to indicate respondents who had never engaged in TU compared to those who had previously used at least one of these tobacco products (no tobacco usage vs. tobacco usage).

#### Alcohol consumption (AC)

Three items were used to measure the prevalence of AC including drunkenness and binge drinking among adolescents [35]. First, respondents were asked “how many days (if any) have you drunk alcohol in your life?”, and provided with responses ranging from 1 (never) to 7 (30 days or more). This was recoded as a dichotomous indicator never drank alcohol vs. drank previously (≥1). Two additional items asked respondents about their experience of drunkenness during the last 30 days (current prevalence) or at any point during their life (lifetime prevalence). For each of these variables responses provided ranged from 1 (never) to 7 (more than 10 times). Two dichotomous variables were thus created, the first indicating no binge drinking vs binge drinking (≥1)), which is a high frequency of AC during the last 30 days, and the second indicating whether the respondent had never been drunk vs drunkenness (≥1) at any time previously.

#### Sleeping habits and patterns (SHPs)

SHPs on school days and weekends/holidays (non-school days) were based on four items asking respondents: “When do you usually go to bed?” and “When do you usually wake up?” Bedtimes ranged in half-hour intervals from “No later than 21:00” to “2:00 or later” for school days, and to “4:00 or later” on non-school days. Waketimes ranged in half-hour intervals from “No later than 5:00” to “8:00 or later” for school days and from “No later than 7:00” to “14:00 or later” on non-school days. Sleep duration for school and non-school days was calculated as the difference between bedtimes and waketimes. For responses at the ends of the scale, we used the minimum/maximum stated time (e.g., 14:00 if the waketime response was “14:00 or later” and 5:00 if the waketime response was “No later than 5:00”). We assessed whether adolescents in the sample slept 9 hours or more based on current international guidelines recommending that school-aged children sleep between 9-11 hours and adolescents 8-10 hours [36, 37].

### Covariates

Socioeconomic circumstances was assessed using (a) *Perceived family wealth*, which is a relative measure of wealth, based on the adolescent’s perception of their family’s economic situation. This was categorised as: (1) quite or very well off, (2) average, (3) not so well off/not at all well off; (b) The *Family Affluence Scale* (FAS) is a widely used and validated absolute measure of wealth [38]. It is scored using six items based on a family’s material possessions and lifestyle including number of: cars (0, 1, 2 or more); bathrooms (0, 1, 2, 3 or more), computers (0, 1, 2, > 2); and holidays abroad during the last 12 months (0, 1, 2, 3 or more); and whether they shared a bedroom (no/yes); or had a dishwasher (no/yes). The responses were categorised into tertiles of low, medium, or high family affluence [30]. *Family situation* described the composition of each household and was categorised to include adolescents residing (a) with both parents; (b) in reconstructed families (including step-parents); (c) single parent households and (d) in foster/children’s home or with other caregivers.*Migrant background* was categorised to indicate respondents who were foreign born, those born in Sweden and the migrant status of their parents, that is, whether one or both parents were born abroad. Finally, sex and the three sampled age groups [30] were used to stratify the analyses.

### Analytical Strategy

First, multiple clusters of HLBs were identified by estimating a series of Latent class analysis (LCA) using PROC LCA command (SAS version 1.3.2) [39]. Five successive sex and age-stratified models of 2 to 6 clusters were estimated separately. One thousand iterations of each model using random starting values were fitted to ensure model identification, missing data was assumed to be at random and handled within the EM algorithm. G^2^ frequencies were compared and the solution with the most common and lowest G^2^ value was identified as reaching a maximum likelihood solution [39]. Statistical fit indices including the Akaike Information Criterion (AIC) [40] and Bayesian Information Criterion (BIC) [41] were examined to determine the number of clusters that best fit the data; low AIC and BIC values are generally deemed to be good fitting models [39]. In addition, the appropriate number of clusters were chosen based on conceptual considerations, theory, cluster distinctiveness and interpretability [39].

The second step of the analysis consisted of conducting multinomial logistic regression to assess the socioeconomic and demographic correlates of each identified cluster while taking into account classification uncertainty in each latent cluster. The likelihood of cluster membership were described as odds ratio (OR) with 95% confidence interval (CI). Analyses were conducted in Stata version 16.1.

## RESULTS

### Descriptive characteristics of the sample

Table 1 shows the descriptive characteristics of the final sample (N = 3 937). The overall distribution was fairly equal by sex (Girls: 51% vs. Boys: 49%). The proportion of respondents increased by age: Approximately 28.2% of the sample were 11-year-olds, 33.8 % were 13-year-olds and 38.0 % were 15-year-olds. Overall, the mean age was 13.2 years (SD = 1.6). There were notable differences in the proportion of adolescents living with both parents (64.8%), compared to those living in reconstructed families (12.9 %), single parent households (7.1%) or foster/children’s home or other caregivers (4.3%). Regarding migrant background, 10.2% of the sample were foreign born whilst the majority were born in Sweden 64.6% and of those born in Sweden 12.7% had one parent with a migrant background and 12.5 % had both parents with a migrant background. The distribution of FAS indicated that approximately 17.5 % of respondents were classified as having high affluence, the majority were classified as having medium affluence (61.7 %) and 17.7% as having low affluence. Regarding perceived family wealth, the majority perceived their family as being quite or very well off (80.7%). Approximately 15% perceived their family as having average wealth and 2.5% as not at all/not so well off.

**Table 1.** Socioeconomic and demographic characteristics of the sample.

|  | N | Percent |
| --- | --- | --- |
| **Sex** |  |  |
| Boys | 1 939 | 49.25 |
| Girls | 1 998 | 50.75 |
| **Age** |  |  |
| 11 | 1 110 | 28.19 |
| 13 | 1 330 | 33.78 |
| 15 | 1 497 | 38.02 |
| **Family situation** |  |  |
| Both parents | 2 551 | 64.80 |
| Reconstructed Family | 507 | 12.88 |
| Single parent | 280 | 7.11 |
| Foster/children's home or other | 169 | 4.29 |
| Missing | 430 | 10.92 |
| **Migrant background** |  |  |
| Swedish born | 2 542 | 64.57 |
| Foreign born | 402 | 10.21 |
| One Parent Migrant | 501 | 12.73 |
| Both Parents Migrants | 492 | 12.50 |
| **Family Affluence Scale (FAS)** |  |  |
| High affluence | 689 | 17.50 |
| Middle affluence | 2 338 | 59.39 |
| Low affluence | 789 | 20.04 |
| Missing | 121 | 3.07 |
| **Perceived family wealth** |  |  |
| Fairly or very well off financially | 3 177 | 80.70 |
| Average financially | 579 | 14.71 |
| Not at all/not so well off | 97 | 2.46 |
| Missing | 84 | 2.13 |

Source: 2017/18 Swedish HBSC study, authors analysis

### Model testing and selection

Analyses for the total dataset indicated that there were better fitting models for identifying distinct clusters by sex and age separately (Full results not shown). Hence, model testing was conducted separately by sex and age, with sequential analysis of nested (constrained and unconstrained) models for measurement invariance (MI). Model fit indices are presented for sex and age separately (Supplementary Table 1). All participants were assigned to the cluster in which they had the highest probability of membership, which were well-identified, interpretable, and conceptually meaningful [42].

#### Comparison of the clusters by sex

We identified four distinct clusters, named according to the HLBs that were most prominent: ***Cluster 1*** *- “Mixed eating behaviours and habits (EBH), physical activity (PA) and low alcohol consumption (AC)”;* ***Cluster 2*** *- “Healthy lifestyle behaviours”;* ***Cluster 3*** *- “Unhealthy lifestyle behaviours”;* ***Cluster 4*** *- “Breakfast, low alcohol consumption (AC) and tobacco usage (TU)”*.The cluster membership probabilities by sex indicated that 6.4% more girls were classified as being in the healthy lifestyle behaviours cluster than boys (21.5 % boys vs 27.9 % girls). In contrast, boys were overrepresented in three remaining clusters: mixed EBH, PA and low AC (31% boys vs 28.4% girls), unhealthy lifestyle behaviours (12.9% boys vs 9.9% girls) and the breakfast and low AC and TU 34.7% boys vs 33.8% girls) cluster.

#### Item-response probabilities and cluster characteristics

Figure 1 shows the item-response probabilities for the model stratified by sex. The item-response probabilities are an indication of the likelihood that respondents engaged in each of the fifteen behaviours. It is the underlying pattern of responses to those behaviours that determine each clusters. The item-response probabilities therefore provide an overall picture of the identified clusters.

**Figure 1.**
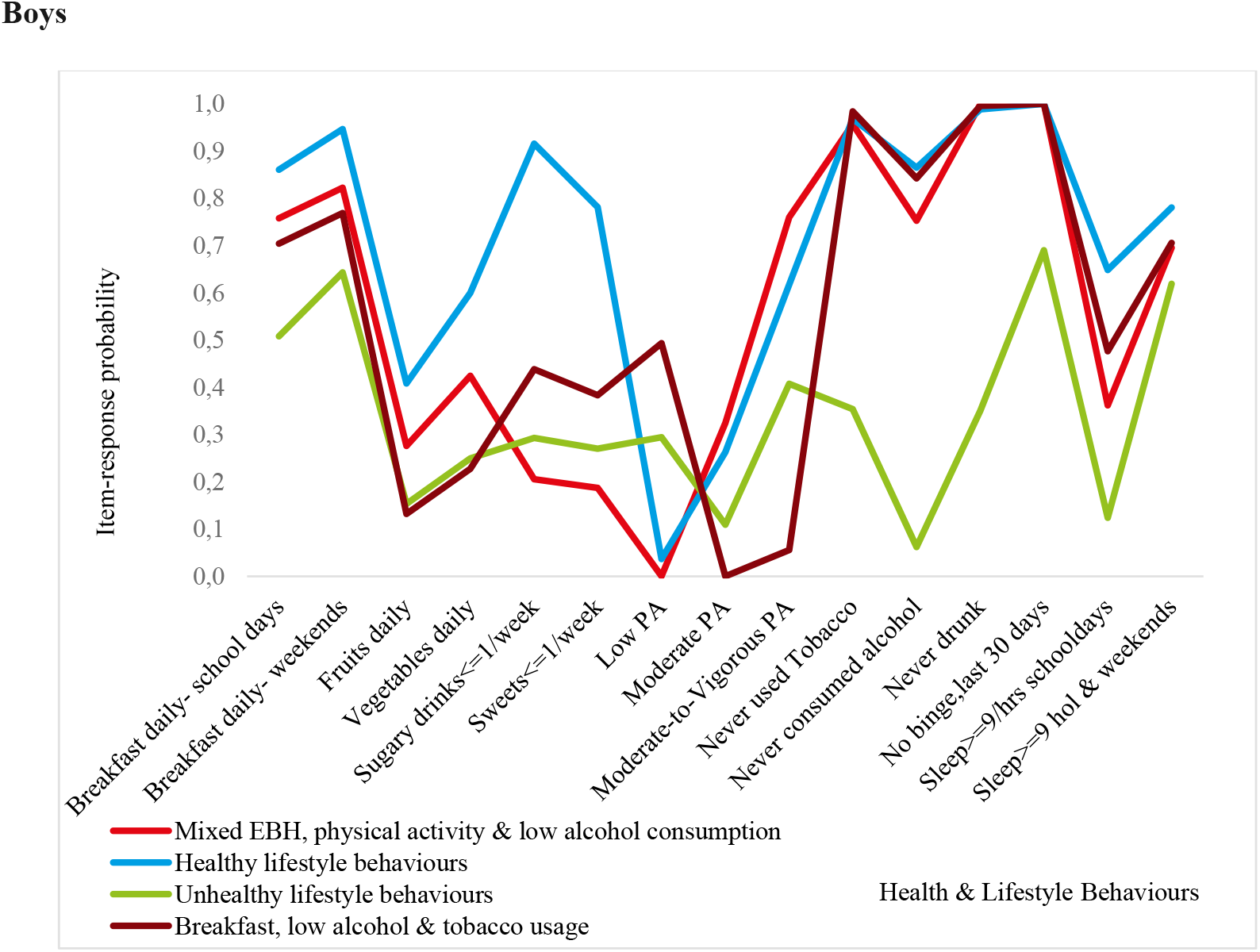
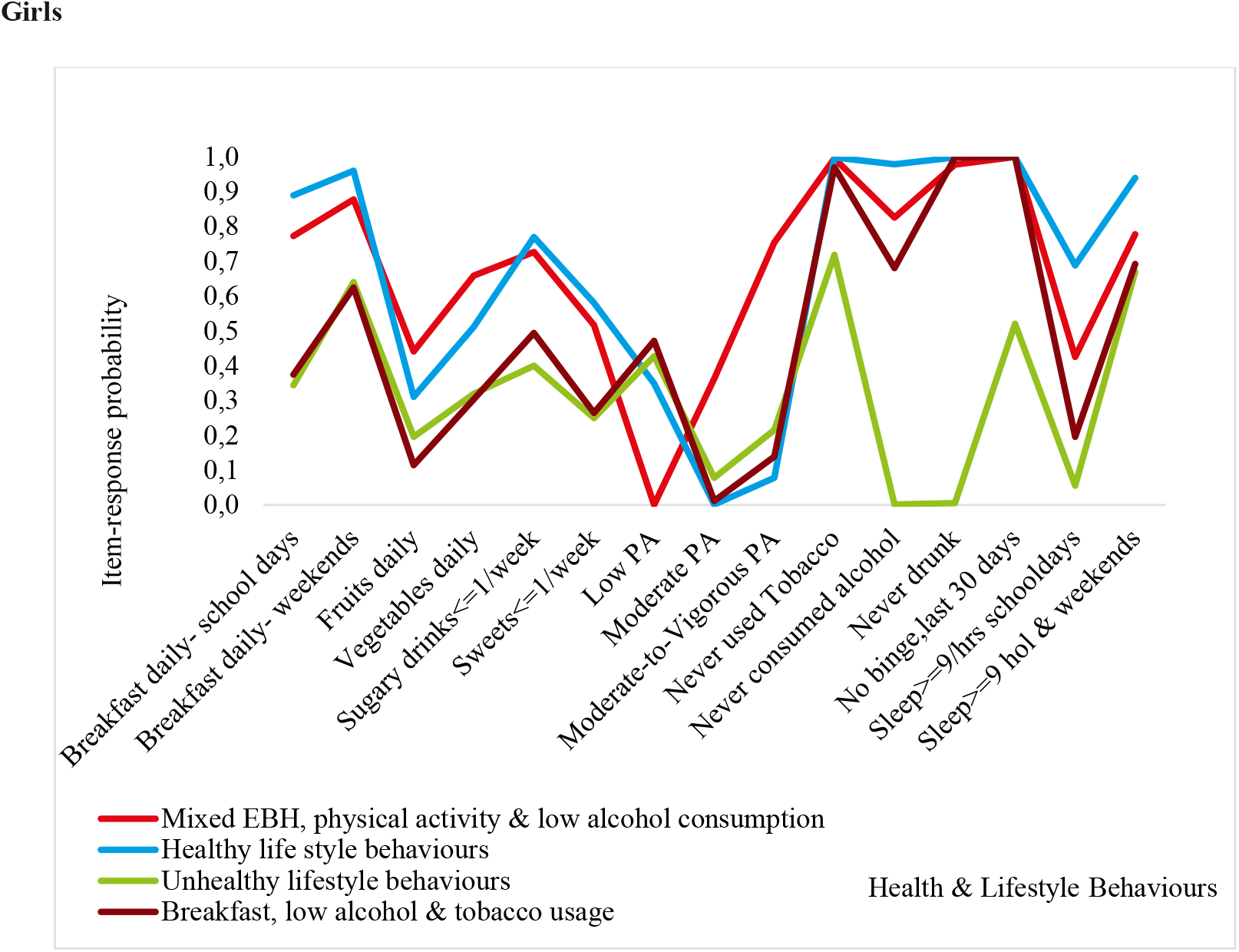
Graphical displays of item-response probabilities across each of the clusters for boys and girls.

The *mixed EBH, PA and low AC cluster* (Cluster 1) was characterised by adolescents with relatively healthy behaviours including: a high probability of eating breakfast, moderate daily consumption of vegetables and meeting the sleep recommendations on non-school. In contrast, they were less likely to have consumed sugary soft drinks, sweets, alcohol or to have used tobacco. In addition, they were less likely to engage in PA school (i.e. low and high PA) but were instead more likely to engage in VPA, that is, sport activities during their leisure time. Whilst the above describes indicators of healthy behaviours, this group had a low probability of consuming fruits on a daily basis or to meet the recommended hours of sleep on school days. As it pertains to breakfast consumption, TU and AC boys and girls engaged in similar HLBs. There were however some differences in the item-response probabilities by sex. Girls had 16% respective 24% higher probability of consuming fruits (28 % boys vs 44 % girls) and vegetables (42 % boys vs 66 % girls) daily. Similarly, the responses indicated that girls had a higher probability of consuming sugary soft drinks once a week or less when compared to boys (21 % boys vs 73 % girls) and sweets once a week or less (19 % boys vs 52 % girls). Indicating lower rates of consumption. The majority of adolescents cluster members did not meet sleep recommendations on school days (36 % boys vs 43 % girls) but they were more likely to do so on weekends/non-school days (70 % boys vs 78 % girls).

Members of the *healthy lifestyle behaviours cluster* (Cluster 2) had a higher likelihood of engaging in positive HLB such as a high probability of eating breakfast both on school days and non-school days, moderate engagement in PA, low probability of having ever used tobacco or consuming alcohol. Furthermore, the majority met sleep recommendations on school days and non-school days. There were however some differences in HLBs by sex related to the daily consumption of fruits (41 % boys vs 31 % girls) and vegetables (60 % boys vs 51 % girls), which was higher among boys than girls. Engagement in PA also varied by sex, where boys were more likely to participate in sports during their leisure time (VPA) and had overall higher PA levels. For example, among boys 62% reported engaging in VPA whilst approximately 26% engaged in high levels of PA. In contrast, girls had a low probability of engaging in VPA (8 %) whilst approximately 35% engaged low levels of PA.

In comparison to the other clusters, the majority of adolescents classified in the *unhealthy lifestyle behaviours cluster* (Cluster 3) were likely to engage in the negative aspects of the five health and lifestyle behaviours included in this study. There are exceptions; this includes, for instance, a relatively high probability of eating breakfast on school days (51% boys vs 64 % girls) and on weekends (64% boys), meeting sleep recommendations on non-school days (62 % boys vs 67 % girls), never engaged in binge drinking (69 % boys vs 52 % girls) and girls were less likely to have ever used tobacco (72%).

Adolescents classified in the breakfast, low AC and TU cluster (Cluster 4) were more likely to engage in unhealthy eating behaviours and low levels of PA compared to the previously discussed clusters. There were also notable differences by sex. For instance, boys had a high probability of consuming breakfast on school days when compared to girls (70 % boys vs 37 % girls). Although the sex differences in the consumption of breakfast was greatly reduced on non-school days (77 % boys vs 62 % girls), consumption among boys remained higher than for girls.

### Comparison of the clusters by age

The results from the LCA age-stratified models indicated that although the four-cluster model had a smaller AIC, the three-cluster model was more interpretable and conceptually meaningful. The three identified clusters based on the most prominent item-response probabilities were: ***Cluster 1*** *-“Unhealthy lifestyle behaviours”;****Cluster 2*** *-“Moderately healthy lifestyle behaviours”;* ***Cluster 3*** *-“Healthy lifestyle behaviours”*. Age differences in cluster membership probabilities show a clear gradient in the proportion of 11-year-olds in each cluster, ranging from 21% in the *unhealthy lifestyle cluster*, 35.2% in the *moderately healthy lifestyle behaviours* cluster and 43.8% in the *healthy lifestyle behaviours cluster*. In contrast, 13-year-olds were on opposite ends of the health behavioural continuum with 29.4%, 16.2% and 54.4% categorised as being in the unhealthy, *moderately healthy and* healthy lifestyle behaviours cluster respectively. Finally, 22% of 15-year-olds were in the *unhealthy lifestyle behaviours cluster*, but the majority were classified in the *moderately healthy behaviours cluster* at 43% whilst approximately 35% were classified in the *healthy lifestyle behaviours cluster*.

### Item-response probabilities and cluster characteristics

Figure 2, shows the item-response probabilities by age. Of the three HLBs identified, the *healthy lifestyle behaviours cluster* (Cluster 1) was characterised by a high engagement in positive HLBs, although this declined with age. EBH remained consistent across age, where approximately 40% respective 60% of this group consumed fruits and vegetables daily; and the majority consumed breakfast. Further, consumption of sugary drinks was similar across all the age groups (approximately 70%) but the probability of eating sweets once a week or less decreased with age, from 62% among 11-year-olds to 47% among 15-year olds. This suggests that the frequency of sweet consumption was higher among older adolescents. In addition, two differences in health behaviours emerged between 11-year-olds and the older age groups. These were related to sleep recommendations on school days and the consumption of alcohol: 15-year-olds had the lowest likelihood of meeting sleep recommendations (26 %), followed by 13-year-olds (56%) when compared to 11-year-olds (77%). 15-year-olds were most likely to have consumed alcohol (36%), whilst only 9% respective 5% 11-year-olds and 13-year-olds had ever consumed alcohol.

**Figure 2.**
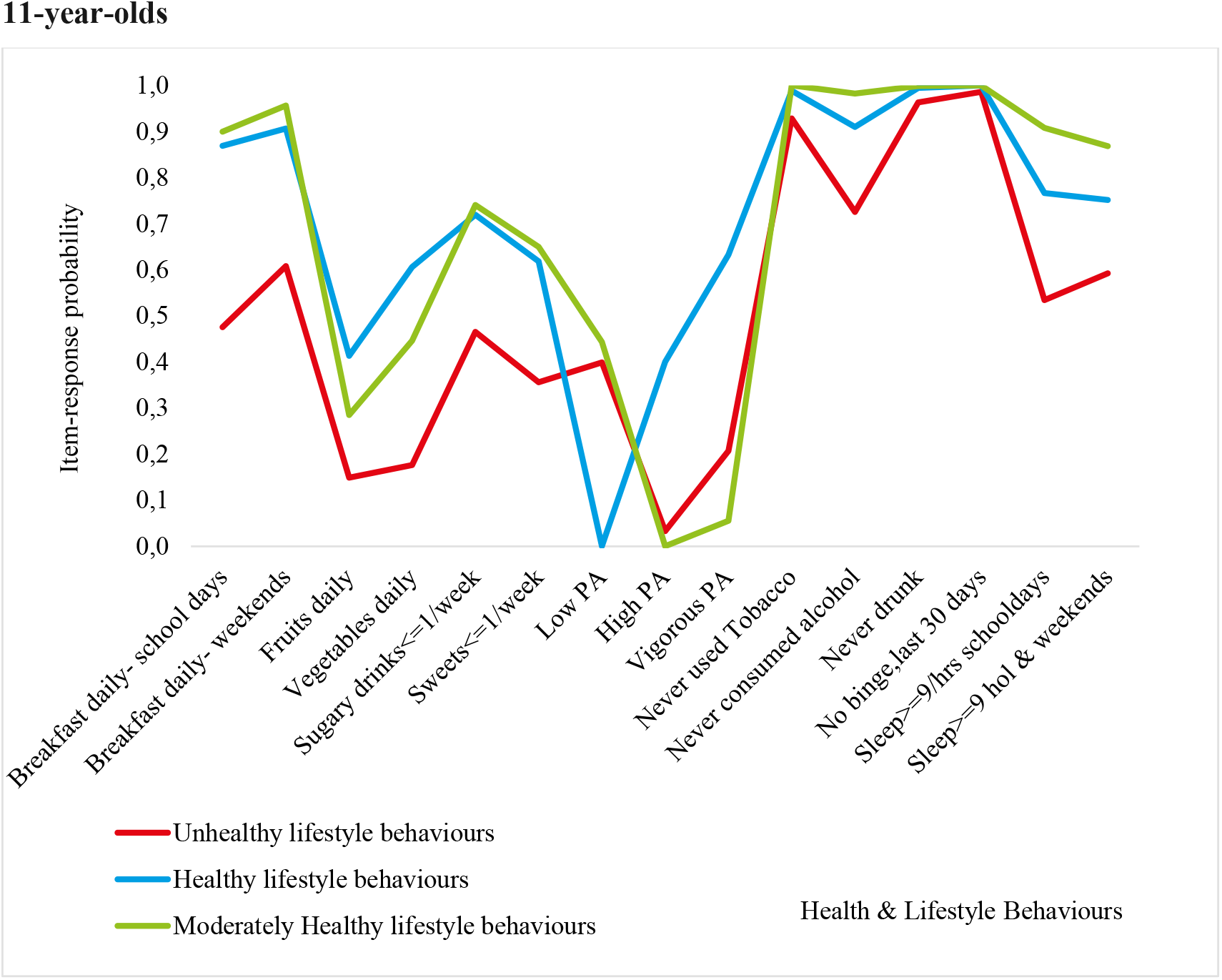
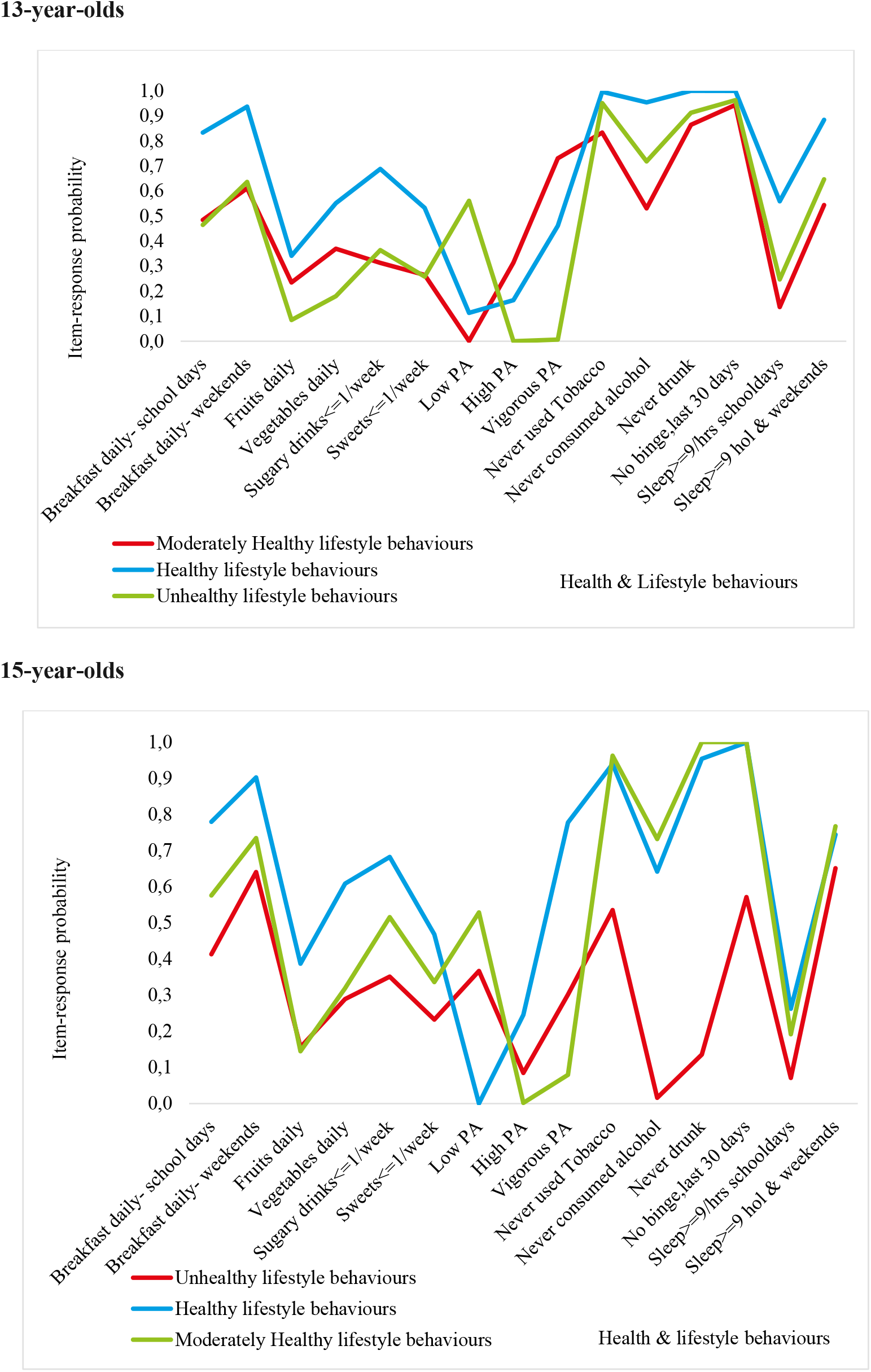
Graphical Displays of Item-Response Probabilities across each of the Age group for 11, 13 and 15-year-olds.

The *moderately healthy lifestyle behaviours* cluster (Cluster 2) was characterised by adolescents with a high probability of consuming breakfast on school days and non-school days, the majority had never engaged in TU or AC. Consumption of fruits and vegetables, and engagement in PA was slightly lower in this cluster compared to the *healthy lifestyle behaviours cluster*. Age differences in health behaviours were also more apparent in this cluster. For instance, compared to 13 and 15-year-olds, more than 90% of 11-year-olds responded positively to eating breakfast on the school days. The consumption of breakfast was however more common among 13 and 15-year-olds (≈ 60%) on weekends. On school days, poor SHPs increased with age. For instance, among 11-year-olds, the likelihood of meeting sleep recommendations were approximately 91%, but 14% respective 19%, among 13 and 15-year-olds.

*The unhealthy behaviours clusters* (Cluster 3) was characterised by poor EBH and PA among adolescents of all ages, but 15-year-olds were most likely to engage in multiple negative HLBs. In addition, across the five HLBs, the probability for healthy engagement was found to be lower compared to the other clusters. For instance, 61% of 11-year-old were likely to eat breakfast on the weekends in this cluster, compared to 91% in the healthy lifestyles cluster. This pattern is similar across the majority of HLBs for 11-year-olds, but engagement in healthy EBH and PA were particularly low, in this cluster. The response pattern for 13-year-olds indicated that they engaged in more healthy behaviours in 6 of the 15 HLBs examined. This includes a low likelihood of TU (83%), AC across the life time (53%), drunkenness (86%) and binge drinking (94%). They were however, more likely to eat breakfast during weekends (61%) and to meet the sleep recommendations on non-school days (54%). The item-response probabilities among 15-year-olds, indicated that they were likely to engage in 11 of 15 HLBs examined. As such, they engaged in more positive HLBs, in relation to four behaviours: breakfast consumption on weekends (64%), TU (54%); binge drinking (57%) and meeting sleep recommendations on non-school days (65 %). They also had a higher probability of engaging in PA.

### Socioeconomic and demographic correlates of cluster membership

The results of the sex-stratified multinomial regression sex-stratified models are shown in Figures 3 (Supplementary table 2). Compared to the *healthy lifestyle behaviours* cluster, common risk factors across all HLB clusters included age, being Swedish born, family situation, medium family affluence and low perceived family wealth. These associations varied by sex and across the HLB clusters. For boys, only age and having medium family affluence was associated with classification in the *Mixed EBH, physical activity and low alcohol consumption* cluster. 15-year-olds girls had 3 times greater risk of classification in the unhealthy behaviours cluster compared to boys of the same age. Furthermore, associations with socioeconomic circumstances were stronger for girls than boys. In particular, girls with medium family affluence and low perceived family wealth were most likely to be classified in the *unhealthy lifestyle behaviours* cluster. Similar risk factors were associated with classification in the *Breakfast, low alcohol and tobacco usage* cluster, but these effects were less pronounced. However, it is worth noting that girls in this cluster were more likely to come from single-parent households and to be foreign-born.

**Figure 3.**
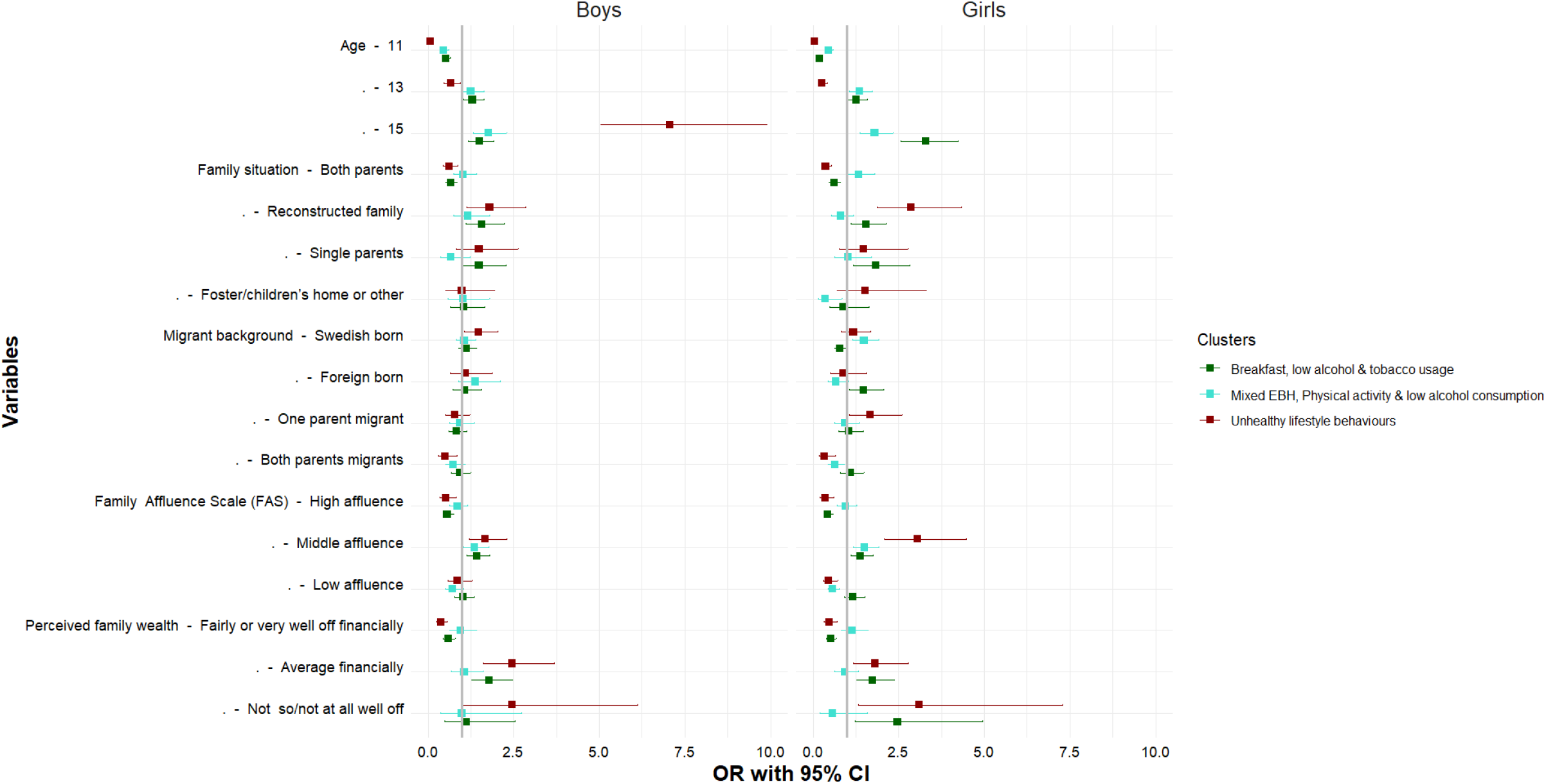
Socioeconomic and demographic correlates of each LCA by sex. Source: Authors analysis, 2017/18 Swedish HBSC study; Notes: Exponentiated coefficients; The reference cluster in the multinomial logistic regression model, the healthy behaviours clusters and the first category of each indicator.

Figure 4 (Supplementary table 3) shows the results of the age-stratified multinomial regression models. The *healthy lifestyle behaviours* cluster is the reference category. We found that girls and adolescents with average perceived family wealth had a significantly higher risk of being classified in the moderately and unhealthy lifestyle behavior clusters across all three age groups Furthermore, being Swedish born among 13-year-olds indicated a higher risks of being classified in the *unhealthy cluster*.The results also showed that 13-year-olds and 15-year-olds from reconstructed families and single parent households had a higher risk of being of classified in the *unhealthy behaviours* cluster. However, unlike the other clusters and age groups, 15-year-olds who were foreign-born, had low family affluence, or had two migrant parents were also more likely to be classified in the unhealthy behavior cluster.

**Figure 4.**
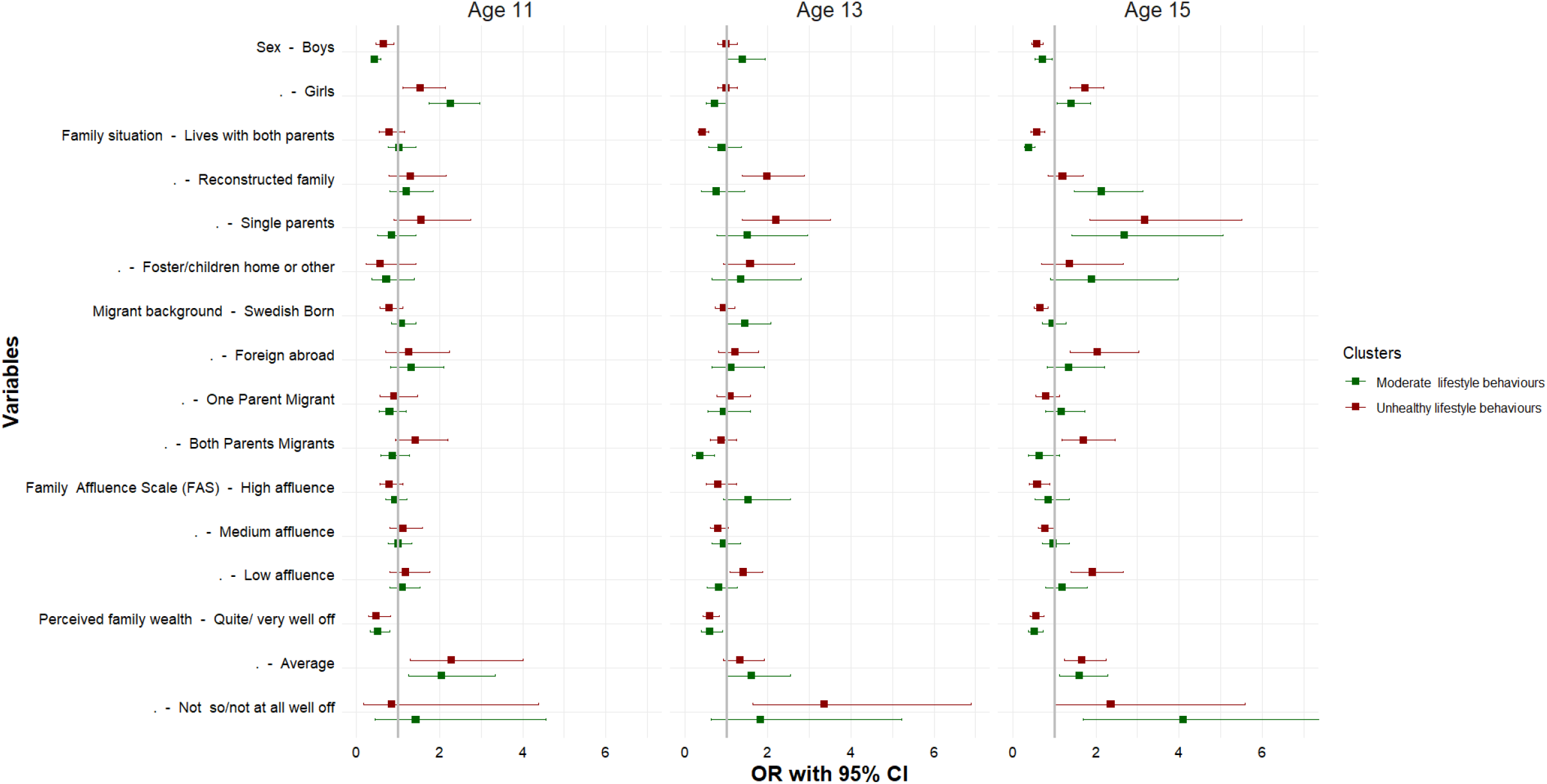
Socioeconomic and demographic correlates of each LCA by age. Source: Authors analysis, 2017/18 Swedish HBSC study; Notes: Exponentiated coefficients; The reference cluster in the multinomial logistic regression model, the healthy behaviours clusters and the first category of each indicator.

## DISCUSSION

Our study revealed distinct health and lifestyle behaviour (HLB) clusters by sex and age separately, rather than for all Swedish children/adolescents.Four distinct HLB clusters were identified for boys and girls in the sex-stratified models, while the age-stratified models pointed to three distinct clusters. However, tests of measurement invariance indicated that the clusters may be meaningfully compared [45]. These results echo the findings from earlier studies [20, 22, 23, 25, 43-45], which suggests that the frequency, intensity and overall HLB patterns are not the same for all groups.

Based on the cluster membership probabilities from the sex-stratified models, we found that boys were overrepresented in every cluster except the healthy behaviours cluster [22, 43, 44]. In contrast to girls, boys were more likely to meet recommendations but less likely to meet other dietary requirements such as the intake of fruits, vegetables, sugar sweetened beverages and sweets. These results differ from at least one study, which found that it was more common for girls to be classified in clusters engaging in poor diet compared to boys [23]. Consistent with prior studies [23, 25, 43], our results also showed that boys consistently reported greater engagement in PA. A recent study by Miranda et al. [46] found that adolescent girls were most likely to be classified in HLB clusters characterised by a high degree of physical inactivity and sedentary behaviours. Meanwhile, our results indicate that AC and TU were more common among boys than girls, especially those classified in the unhealthy behaviours cluster. SHP also varied by sex and across all the clusters.

A similar assessment of cluster membership from the age-stratified models revealed a gradient in HLBs among 11-year-olds and 15-year-olds. For instance, among 11-year-olds, the highest proportion of the sample were classified in the healthy behaviours cluster and the lowest in the unhealthy cluster. In contrast, far less 15-year-olds were classified in the healthy behaviours cluster, instead they were significantly overrepresented in moderately healthy behaviours cluster. The results demonstrated however that a higher proportion of 15-year-olds were classified in the unhealthy behaviours cluster compared to 11-year-olds, but this was slightly lower than for 13-year-olds. In comparison to the other age groups, a greater proportion of 13-year-olds were classified in the unhealthy respective the healthy behaviours cluster. These results are somewhat unexpected, because one would expect that the results for 13-year-olds would be somewhere in between the results for 11- and 15-year olds. A systematic review by Leech and colleagues [43], which included 18 individual studies investigating the clustering of multiple health behaviours found that older children/adolescents tended to comprise clusters with less healthy behaviours [43].

The second aim was to assess the association between socioeconomic and demographic factors and the identified HLBs clusters. Although,we observed that the correlates of cluster membership was similar for boys and girls, a key difference emerged. The association between socioeconomic and demographic indicators and unhealthy behaviour for both boys and girls was strong [46], but slightly stronger for girls. Age and perceived family wealth were consistent predictors of cluster membership. Socioeconomic disadvantage was also found to be consistent predictor, but the relationship varied depending on the indicator. Although FAS was correlated with cluster membership, adolescents’ perception of their family’s wealth was found to be a more consistent predictor of cluster membership. It important to note that the clustering of HLBs are likely to be attenuated or exacerbated by socioeconomic and demographic factors. Prior studies have shown that poor health behaviours coupled with poor socioeconomic circumstances can have a more deleterious health impact, which may increase the risk of morbidity and mortality [3, 16, 22, 24, 43]. For example, there is evidence that adolescents who are physically inactive are at greater risk for both short-and long term health problems [3, 16].

In addition, we found an inverse relationship between socioeconomic circumstances and cluster membership classification, with adolescents from disadvantaged backgrounds being more likely to be classified in the unhealthy behaviours cluster [45, 46]. The association was stronger for family situation such as reconstructed families and single parent households. Overall, there was a strong likelihood for girls, adolescents born abroad, those with two parents born abroad to be classified in the unhealthy behaviours cluster [45, 46]. The likelihood of being classified as unhealthy increased for 11-year-olds and the 15-year-olds. In contrast, 13-year-olds were more likely to belong to both the unhealthy and the healthy clusters compared to the 11-year-olds and the 15-year-olds.

### Strengths and limitations

This study has some limitations. First, the key measures used in the analyses were based on self-reported measures with different reference periods, and they may therefore be subject to recall bias. Second, LCA models have a number of fit indices, which vary based on different distributional assumptions. However, the use of multiple indices allows for a more balanced assessment of the models. Moreover, the results of LCA models were difficult to compare across studies given that the identified clusters were determined by the set of items included in specific models and the operationalisation of these items which vary widely across studies [22, 24, 25, 43, 44].

Despite the limitations, a strength of the current study was the use of self-reported data, collected from children and adolescents in the HBSC study; providing their unique views and experiences on living conditions and health behaviours. Another strength was the use of a relatively large and nationally representative sample, which allowed for group level assessments and differentiation of health and lifestyle behaviours using LCA methods. An advantage of LCA is the possibility to examine the inter/intra-item relationships between multiple health behaviours simultaneously, and to assess numerous combinations of unobserved behaviour patterns before reducing the data into smaller clusters [47]. This current analyses therefore contributes to our understanding of the interlinkage between various health behaviours while allowing for assessments of the differences in health behaviours by factors such as age and sex. Moreover, the use of LCA contributes to the growing scientific literature suggesting that health behaviours are not randomly distributed, but rather cluster within individuals and across similar socioeconomic and demographic characteristics [19, 24, 25, 28].

## CONCLUSION

Adolescence is a critical period of development where lifelong health and behavioural practices are normalised. Many policies and programmes that aim to improve health narrowly focus on specific lifestyle behaviours such as physical activity or eating habits. Our findings suggest that a broader range of health and lifestyle behaviours should be tackled simultaneously, rather than focusing on specific individual/single activities. Given that patterns of health behaviours vary by sex and age, a cluster-based approach,which captures the intersectionality between socioeconomic determinants of health and health behaviours, could lead to more effective and targeted behaviour change models.Our results suggest that it may be prudent to tailor health-promoting interventions to target desired behaviour changes and improvements for boys and girls at different ages. In conclusion, using a multi-behavioral change approach in prevention strategies can help identify and target vulnerable and at-risk adolescents.

## Data Availability

Data requests should be submitted to

## REFERENCE

1. Flodmark, C.-E., Prevention models of childhood obesity in Sweden. Obesity facts, 2018. 11(3): p. 257–262.

2. Neovius, M., J. Sundström, and F. Rasmussen, Combined effects of overweight and smoking in late adolescence on subsequent mortality: nationwide cohort study. Bmj, 2009. 338.

3. Lindberg, L., et al., Association of childhood obesity with risk of early all-cause and cause-specific mortality: A Swedish prospective cohort study. PLoS medicine, 2020. 17(3): p. e1003078.

4. Alfonsi, V., et al., Later school start time: the impact of sleep on academic performance and health in the adolescent population. International journal of environmental research and public health, 2020. 17(7): p. 2574.

5. Vist, G.E., et al., Are the health risks of moist oral snuff (snus) underestimated? Tidsskr Nor Laegeforen, 2020. 140(9).

6. Sweden, T.P.H.A.o. Overweight and obesity among school children 11–15-year-old continues to increase. 2020 2022-05-21]; 20020:[Available from: https://www.folkhalsomyndigheten.se/publicerat-material/publikationsarkiv/o/overweight-and-obesity-among-school-children-1115-year-old-continues-to-increase/.

7. och Livsmedelsverket, F., Förslag till åtgärder för ett stärkt, långsiktigt arbete för att främja hälsa relaterad till matvanor och fysisk aktivitet. Stockholm och Uppsala: Folkhälsomyndigheten och Livsmedelsverket, 2017.

8. Winpenny, E.M., et al., Changes in diet through adolescence and early adulthood: longitudinal trajectories and association with key life transitions. International Journal of Behavioral Nutrition and Physical Activity, 2018. 15(1): p. 1–9.

9. Taverno Ross, S.E., et al., Longitudinal changes in physical activity and sedentary behavior from adolescence to adulthood: comparing U.S.-born and foreign-born populations. Journal of physical activity & health, 2014. 11(3): p. 519–527.

10. Aira, T., et al., Physical activity from adolescence to young adulthood: patterns of change, and their associations with activity domains and sedentary time. International Journal of Behavioral Nutrition and Physical Activity, 2021. 18(1): p. 1–11.

11. Lien, N., L.A. Lytle, and K.-I. Klepp, Stability in consumption of fruit, vegetables, and sugary foods in a cohort from age 14 to age 21. Preventive medicine, 2001. 33(3): p. 217–226.

12. Reitsma, M.B., et al., Spatial, temporal, and demographic patterns in prevalence of smoking tobacco use and initiation among young people in 204 countries and territories, 1990–2019. The Lancet Public Health, 2021. 6(7): p. e472–e481.

13. Hedin, G., et al., Insomnia in relation to academic performance, self-reported health, physical activity, and substance use among adolescents. International Journal of Environmental Research and Public Health, 2020. 17(17): p. 6433.

14. Raffetti, E., et al., Longitudinal association between tobacco use and the onset of depressive symptoms among Swedish adolescents: the Kupol cohort study. European Child & Adolescent Psychiatry, 2019. 28(5): p. 695–704.

15. Tjora, T., J.C. Skogen, and B. Sivertsen, Establishing the association between snus use and mental health problems: A study of Norwegian college and university students. Nicotine Tob Res, 2022.

16. Murray, C.J., et al., Global burden of 87 risk factors in 204 countries and territories, 1990– 2019: a systematic analysis for the Global Burden of Disease Study 2019. The Lancet, 2020. 396(10258): p. 1223–1249.

17. Organization, W.H., Global action plan for the prevention and control of noncommunicable diseases 2013-2020. 2013, World Health Organization.

18. Andersson, E., K. Welin, and K. Carlsson, Kostnader för fetma i Sverige idag och år 2030. IHE Rapport, 2018. 3.

19. Prochaska, J.O., Multiple health behavior research represents the future of preventive medicine. Preventive medicine, 2008. 46(3): p. 281–285.

20. Ahmad, K., et al., Clustering of Lifestyle and Health Behaviours in Australian Children and Their Relationship with Obesity, Self-Rated Health and Quality of Life. 2022.

21. Poortinga, W., The prevalence and clustering of four major lifestyle risk factors in an English adult population. Preventive Medicine, 2007. 44(2): p. 124–128.

22. D’Souza, N.J., et al., A systematic review of lifestyle patterns and their association with adiposity in children aged 5–12 years. Obesity Reviews, 2020. 21(8): p. e13029.

23. Nolan, A. and E. Smyth, Clusters of health behaviours among young adults in Ireland. 2020: Research Series.

24. Parker, K.E., et al., Activity-related behavior typologies in youth: a systematic review. International Journal of Behavioral Nutrition and Physical Activity, 2019. 16(1): p. 1–13.

25. Miranda, V.P.N., et al., Use of latent class analysis as a method of assessing the physical activity level, sedentary behavior and nutritional habit in the adolescents’ lifestyle: A scoping review. PloS one, 2021. 16(8): p. e0256069.

26. Degerlund Maldi, K., et al., Widespread and widely widening? Examining absolute socioeconomic health inequalities in northern Sweden across twelve health indicators. International Journal for Equity in Health, 2019. 18(1): p. 1–12.

27. Lundberg, O., The next step towards more equity in health in Sweden: how can we close the gap in a generation? Scandinavian journal of public health, 2018. 46(22_suppl): p. 19–27.

28. Prochaska, J.J., B. Spring, and C.R. Nigg, Multiple health behavior change research: an introduction and overview. Preventive medicine, 2008. 46(3): p. 181–188.

29. Currie, C., S. Nic Gabhainn, and E. Godeau, The Health Behaviour in School-aged Children: WHO Collaborative Cross-National (HBSC) study: origins, concept, history and development 1982–2008. International journal of public health, 2009. 54(2): p. 131–139.

30. Inchley, J., et al., Health Behaviour in School-aged Children (HBSC) study protocol: Background, methodology and mandatory items for the 2017/18 survey. St Andrews: CAHRU, 2018.

31. Nardone, P., et al., Dietary habits among Italian adolescents and their relation to socio-demographic characteristics. Ann Ist Super Sanità, 2020. 56(4): p. 504–513.

32. Hagström, C., Nu är det fredagsmys! Chips och gemenskap när vardag blir helg. Budkavlen: Tidskrift för etnologi och folkloristik, 2012. 91: p. 9–25.

33. Organization, W.H., Global recommendations on physical activity for health. Geneva; 2010. 2017.

34. Folkhälsomyndigheten, F., Skolbarns hälsovanor i Sverige 2017/18. 2018.

35. Inchley, J., et al., Adolescent alcohol-related behaviours: trends and inequalities in the WHO European Region, 2002–2014: observations from the Health Behaviour in School-aged Children (HBSC) WHO collaborative cross-national study. 2018: World Health Organization. Regional Office for Europe.

36. Hirshkowitz, M., et al., National Sleep Foundation’s updated sleep duration recommendations: final report. Sleep Health, 2015. 1(4): p. 233–243.

37. Gariepy, G., et al., How Are Adolescents Sleeping? Adolescent Sleep Patterns and Sociodemographic Differences in 24 European and North American Countries. Journal of Adolescent Health, 2020. 66(6, Supplement): p. S81–S88.

38. Corell, M., et al., Does the family affluence scale reflect actual parental earned income, level of education and occupational status? A validation study using register data in Sweden. BMC Public Health, 2021. 21(1): p. 1–11.

39. Lanza, S.T., et al., Proc LCA & Proc LTA users’ guide (Version 1.3. 2). University Park: The Methodology Center, Penn State, 2015.

40. Akaike, H., Factor analysis and AIC, in Selected papers of hirotugu akaike. 1987, Springer. p. 371–386.

41. Raftery, A.E., Bayesian model selection in social research. Sociological methodology, 1995: p. 111–163.

42. Russell Jonsson, K., et al., Health behaviors and subsequent mental health problems during the COVID-19 pandemic: A longitudinal analysis of adults in the UK. Frontiers in Public Health, 2023. 10.

43. Leech, R.M., S.A. McNaughton, and A. Timperio, The clustering of diet, physical activity and sedentary behavior in children and adolescents: a review. International Journal of Behavioral Nutrition and Physical Activity, 2014. 11(1): p. 1–9.

44. Cabanas-Sánchez, V., et al., Association between clustering of lifestyle behaviors and health-related physical fitness in youth: the UP&DOWN study. The Journal of pediatrics, 2018. 199: p. 41-48. e1.

45. Paulsson Do, U., et al., Vulnerability to unhealthy behaviours across different age groups in Swedish Adolescents: a cross-sectional study. Health Psychology and Behavioral Medicine, 2014. 2(1): p. 296–313.

46. Carlerby, H., et al., Risk behaviour, parental background, and wealth: A cluster analysis among Swedish boys and girls in the HBSC study. Scandinavian Journal of Public Health, 2012. 40(4): p. 368–376.

47. Lanza, S.T. and B.L. Rhoades, Latent class analysis: an alternative perspective on subgroup analysis in prevention and treatment. Prevention science, 2013. 14(2): p. 157–168.

